# Optimized patient-specific immune checkpoint inhibitors therapy for cancer treatment based on tumor immune microenvironment modeling

**DOI:** 10.1101/2024.04.08.24305526

**Authors:** Yao Yao, Frank Youhua Chen, Qingpeng Zhang

**Author notes:** Corresponding author: Qingpeng Zhang.

## Abstract

**Objective:** Enhancing patient response to immune checkpoint inhibitors (ICIs) is crucial in cancer immunotherapy. We aim to create a data-driven mathematical model of the tumor immune microenvironment (TIME) and utilize deep reinforcement learning (DRL) to optimize patient-specific ICI therapy combined with chemotherapy (ICC).

**Methods:** Using patients’ genomic and transcriptomic data, we develop an ordinary differential equations (ODEs)-based TIME model to characterize interactions among chemotherapy, ICIs, immune cells, and cancer cells. A DRL algorithm is trained to determine the personalized optimal ICC therapy.

**Results:** Numerical experiments with real-world data demonstrates that the proposed TIME model can predict ICI therapy response. The DRL-derived personalized ICC therapy outperforms predefined fixed schedules. For tumors with extremely low CD8+T cell infiltration (“extremely cold tumors”), DRL recommends high-dosage chemotherapy alone. For tumors with higher CD8+T cell infiltration (“cold” and “hot tumors”), an appropriate chemotherapy dosage induces CD8+T cell proliferation, enhancing ICI therapy outcomes. Specifically, for “hot tumors,” chemotherapy and ICI are administered simultaneously, while for “cold tumors,” a mid-dosage of chemotherapy makes the TIME “hotter” before ICI administration. However, a number of “cold tumors” with rapid resistant cancer cell growth, ICC eventually fails.

**Conclusion:** This study highlights the potential of utilizing real-world clinical data and DRL to develop personalized optimal ICC by understanding the complex biological dynamics of a patient’s TIME. Our ODE-based TIME model offers a theoretical framework for determining the best use of ICI, and the proposed DRL model may guide personalized ICC schedules.

**SIGNIFICANCE STATEMENT:** Our research presents a novel data-driven approach to personalized cancer treatment by combining artificial intelligence and mathematical models of the tumor’s surrounding environment, known as the tumor immune microenvironment (TIME). This innovative method allows for the optimization of patient-specific immune checkpoint inhibitors and combined chemotherapy therapy. By utilizing deep reinforcement learning, our approach can adapt and improve treatment strategies for individual patients, ultimately maximizing the effectiveness of cancer therapies. This pioneering work has the potential to significantly enhance clinical decision-making and improve patient outcomes, paving the way for personalized cancer immunotherapy.

## INTRODUCTION

Immune checkpoint inhibitors (ICIs) have significantly advanced the treatment of various cancers, including lung cancer, melanoma, and head and neck cancers.^1,2^ These innovative therapies target the immune checkpoints (ICs), cellular mechanisms that tumor cells exploit to suppress the activity of activated T cells and thus evade immune surveillance. However, patients’ response rate to ICI is notably low.^3^ Research into Tumor Immunity in the Microenvironment (TIME) has identified that lack of tumor-infiltrating lymphocytes (TILs) accounts for most non-responders to ICI therapy.^4^ These insufficiently inflamed tumors, cold tumors, are the focus of efforts to improve treatment effectiveness.^5^ Researchers have attempted to combine chemotherapy and ICI to yield better synergetic treatment results based on the knowledge that chemotherapy can promote immunity.^6–8^ First, chemotherapy can cause tumor debulking, which can alleviate immune suppression exerted by tumor masses.^9^ Second, chemotherapy can induce a type of tumor cell death called immunogenic apoptosis, which can stimulate the T cell inflammation.^6,10^ Third, certain chemotherapy drugs can modulate the immune ecosystem by depleting immune suppressive cells like regulatory T cells (Tregs) and myeloid-derived suppressor cells (MDSCs) while promoting recruitment and activation of dendritic cells (DCs).^11–14^ Clinical trials of ICI and chemotherapy combination (ICC) have yielded promising outcomes, such as improved overall response rates (ORR) and progression-free survival (PFS) in various cancer types.^15–17^ However, the results across trials have been inconsistent, and the underlying reasons for the successes and failures remain elusive. A particular concern is that chemotherapy delivered at or near the maximum-tolerated dosage (MTD) could impair anti-tumor immunity, potentially undermining the benefits of ICI.^18^ The optimal dosage of chemotherapy for ICC remains poorly understood, which hinders our ability to tailor the dynamic evolution of the TIME for personalized ICC therapy.

To fully unleash the synergetic potential potency of chemotherapy and ICI combination, two questions remained to be addressed:^6^ (a) Should chemotherapy be used at an MTD or an appropriate dosage for individual patients? (b) What sequence should ICI and chemotherapy be administrated in? What mechanism results in the difference in the efficacies among different sequences? Therefore, we aim to answer these two questions, especially in the context of patient heterogeneity.

The traditional way to test the safety and efficacy of pharmaceutical therapy is via clinical trials. However, clinical trials are expensive, and two drug combinations at different dosages and administration schedules are enormous. ^19^ It was pointed out that the preclinical rationale of ICC is insufficient, often resulting in failed clinical trials.^6^ Consequently, computer modeling is a valuable tool to explore the mechanism of drug actions (MODA) and predict the outcome of individualized cancer therapy.^20,21^ Various mathematical models, including ordinary differential equations (ODEs) and partial differential equations (PDEs), have been applied to depict tumor growth, tumor-immune system interaction, and treatment intervention.^22,23^ Based on these models’ prediction of the TIME evolution, researchers further applied optimization techniques to derive the optimized therapeutic regimes. ^24–26^

De pillis modeled the dynamics of the tumor cells and immune cells in a system of ODEs and simulated the combined treatment results of chemotherapy and cytokine interleukin-2 (IL-2).^22^ Since then, new medical mechanisms and treatments have been modeled in the forms of ODEs, for example, regulatory T cells.^25^ In the recent three years, the mechanisms of ICIs were modeled to predict the treatment result of ICI monotherapy or ICI combined with other therapies, for example, radiotherapy and androgen deprivation therapy (ADT).^27–29^ However, these models have not extended to the interplay between ICI and chemotherapy. From the perspective of parameter estimation, most models adopted parameters from literature and hypotheses, with a few models calibrated by patient-specific data such as computed tomography (CT).^30^ The information on genomic and transcriptomic features has been integrated into the clinical model to describe ICI treatment outcomes. However, these clinical models can not provide mechanical reasoning as mathematical ones.^31^ Furthermore, despite the success in predicting treatment efficacy, models for optimal therapeutic regimes are under-researched. Existing optimal control approaches fell short in modeling the inherent complexity of the ICC mechanism in the dynamic TIME.^25^ Reinforcement learning (RL), an advanced optimization tool specialized in complicated and extensive Markov decision processes (MDP), is well-suited to address problems that require achieving the maximum long-term reward for individual patients.^32^

In this paper, to the best of our knowledge, we present the first mathematical model to explore the dynamic evolution of the TIME during ICC therapy by characterizing the interplay between ICIs and chemotherapy. Our model is calibrated by the biological and clinical knowledge from the latest literature, including the proliferation and suppression of CD8^+^T cells, immune checkpoints’ suppression of CD8^+^T cells’ functions, and the synergy and impairment effects of chemotherapy on the immune system. Then, we use data, including genomic and transcriptomic features, in the TCGA SKCM (Skin Cutaneous Melanoma study in The Cancer Genome Atlas Program) dataset and Liu dataset to estimate the parameters indicating TIME components interaction in our models.^31,33^ Finally, we use a TCGA SKCM patient cohort to train a Deep Reinforcement Learning (DRL) agent to derive the personalized optimal ICC therapy. We found that the DRL-derived policy outperformed baselines and provided novel insights into how ICC steers dynamic TIME evolution and how we may use this steering power to achieve the optimal treatment outcome for individual patients.

## MATERIALS AND METHODS

### TIME dynamical evolutionary model

We denote the kinetics of sensitive tumor cells, resistant tumor cells, CD8^+^T cells, immune checkpoint inhibitor dosage, and the chemotherapy dosage as *T*_*s*_, *T*_*r*_, *L, I*, and *M*, respectively.

### Tumor cells

The treatment efficacy of chemotherapy decays because of the evolutionary resistance. The chemotherapy drug’s elimination of sensitive tumor cells will provide living space for resistant tumor cells. Following Zhang et al.’s work, we use Lotka-Volterra (LV) competition equations to model the growth and interplay of sensitive and resistant tumor cells.^34^

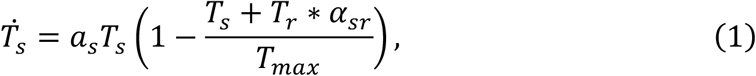

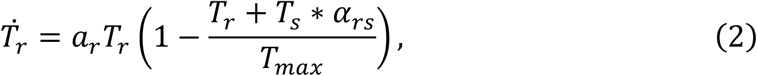

Here, we denote the environment maximal carry capacity of tumor cells as *T*_*max*_, by which the growth of tumor cells is limited. The intrinsic growth rates of sensitive and resistant tumor cells are *a*_*s*_ and *a*_*r*_. The competition effects between sensitive and resistant tumors are represented by *α*_*sr*_ and *α*_*rs*_. The higher *α*_*sr*_ indicates the more substantial competition effects of resistant tumor cells on sensitive ones. The total tumor cell population is *T, T = T*_*s*_ + *T*_*r*_.

### Regulatory T cells

Tumor cells express indoleamine-pyrrole 2,3-dioxygenase (IDO) to promote the accumulation of regulatory T cells.^35^ The regulatory T cells release granzyme to promote the lyse of CD8^+^T cells.^36^ It is the critical factor in deficient T-cell homing to the tumor bed and has been found to contribute to ICI’s clinical failure.^37,38^ Because the ratio of regulatory T cells to CD8^+^T cells (*η*_*RL*_) can be estimated by CIBERSORT based on tissue gene expression profiles (GEPs), instead of modeling the kinetics of regulatory T cells, we directly model this regulation effect proportional to the ratio of regulatory T cells to CD8^+^T cells (*η*_*RL*_) with a regulation rate *h*.^39^

### CD8^+^T cells

There are two pathways determining the proliferation of CD8^+^T cells.^5,40^ First, the presence of antigens comes from three sources: the surface of tumor cells, the immunogenic tumor cell death induced by CD8^+^T cells, and the immunogenic tumor cell death induced by chemotherapy drugs.^41,42^ Similarly, we use the Michaelis-Menten term to model the antigen growth rates out of these three sources: *j*_*C*_*T/*(*k*_*C*_ + *T*), *j*_*L*_*D*_*L*_*T/*(*k*_*L*_ + *D*_*L*_*T*), and *j*_*M*_*D*_*M*_*T*_*s*_*/*(*k*_*M*_ + *D*_*M*_*T*_*s*_), respectively.^43^ Here, *D*_*L*_ and *D*_*M*_ are killing fraction of CD8^+^T cells on tumor cells and chemotherapy drugs on sensitive tumor cells. *j*_*C*_, *j*_*L*_, *j*_*M*_ denote the limiting rate of antigen presence on the surface of tumor cells, released by CD8^+^T cells-induced tumor cell death and released by chemotherapy-induced tumor cell death, respectively. Moreover, *k*_*C*_, *k*_*L*_,*k*_*M*_ are corresponding Michaelis constants, which measure the substrate concentration to produce effects, as when the tumor cell population numerically equals the Michaelis constant, the antigen presence rate is half of *j*_*C*_, *j*_*L*_, *j*_*M*_. As we can perform ssGSEA of MHC-I and MHC-II (two primary classes of major histocompatibility complex molecules) associated HLA genes, we associate the normalized ssGSEA score (*ϕ*) with *j*_*C*_ by the term, 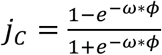. Here, *ω* is the parameter. Second, the patient-specific ability to process antigens. This function is related to the presence of antigen process cells (APCs) and the maturation of APCs. We modeled this ability as *r*. CD8^+^T cells are assumed to die constantly, *m*_*L*_. As we mentioned, the CD8^+^T cells are suppressed by regulatory T cells at a rate of *hη*_*RL*_.

Additionally, tumor cells can directly suppress CD8^+^T cells by secreting FasL, modeled by another Michaelis-Menten term, *−qLT/*(*T* + *u*), where *q* denotes the suppression effects of tumor cells on CD8^+^T cells.

To sum up, the net growth term of CD8^+^T cells is shown below.

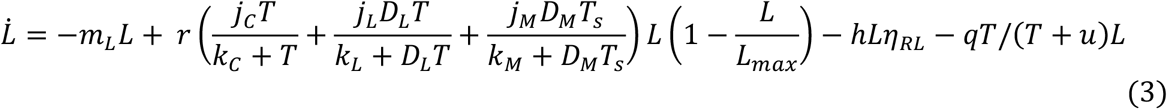

Next, we explain how we model the CD8^+^T cell’s killing effect on tumor cells, *D*_*L*_. We take the fractional cell kill term from literature, 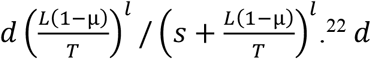 indicates the killing power of CD8^+^T cells. Here, we add a parameter, µ, representing the fraction of CD8^+^T cells suppressed by the checkpoints.

### ICIs effects

ICIs are anti-checkpoint therapy, releasing CD8+T cells suppressed by the immune checkpoints (ICs) and recovering their killing potency. In our model, the original fraction of CD8^+^T cells suppressed by ICs is denoted as µ_0_. The ICIs decrease the fraction to µ_0_(1 *− I*), where *I* is the concentration of ICIs, which decays exponentially with rate *γ*_I_.

### Chemotherapy’s effects

Chemotherapy drug kills all the cells in the ecosystem, *K*_*i*_(1 *− e*^*™M*^)*i, i ∈* {*T*_*s*_, *L*}, where (1 *− e*^*™M*^) is a saturation term, and *K*_*t*_, *i ∈* {*T*_*s*_, *L*} indicate the killing effects on sensitive tumor cells and CD8^+^T cells, respectively. It has been found that most chemotherapy drugs upregulate the PD-L1 expression.^44^ We modeled this effect by the term *p*(1 *− e*^*™M*^), where *p* is the upregulation rate of chemotherapy drugs on checkpoint inhibitors. Similarly, we assumed the concentration of chemotherapy drug decays exponentially with rate *γ*_*M*_.

### ODE model

With all the specific terms depicting the cell’s interactive activities, we derive the following ODE-based TIME dynamic evolutionary model characterizing the interplays between tumor cells, immune cells, and drugs. Their relationships are also depicted in Figure 1. A patient’s TIME state at time t is denoted as 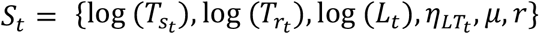. Here, 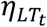 is the ratio of CD8^+^T cell population to tumor cell population.

**Figure 1.**
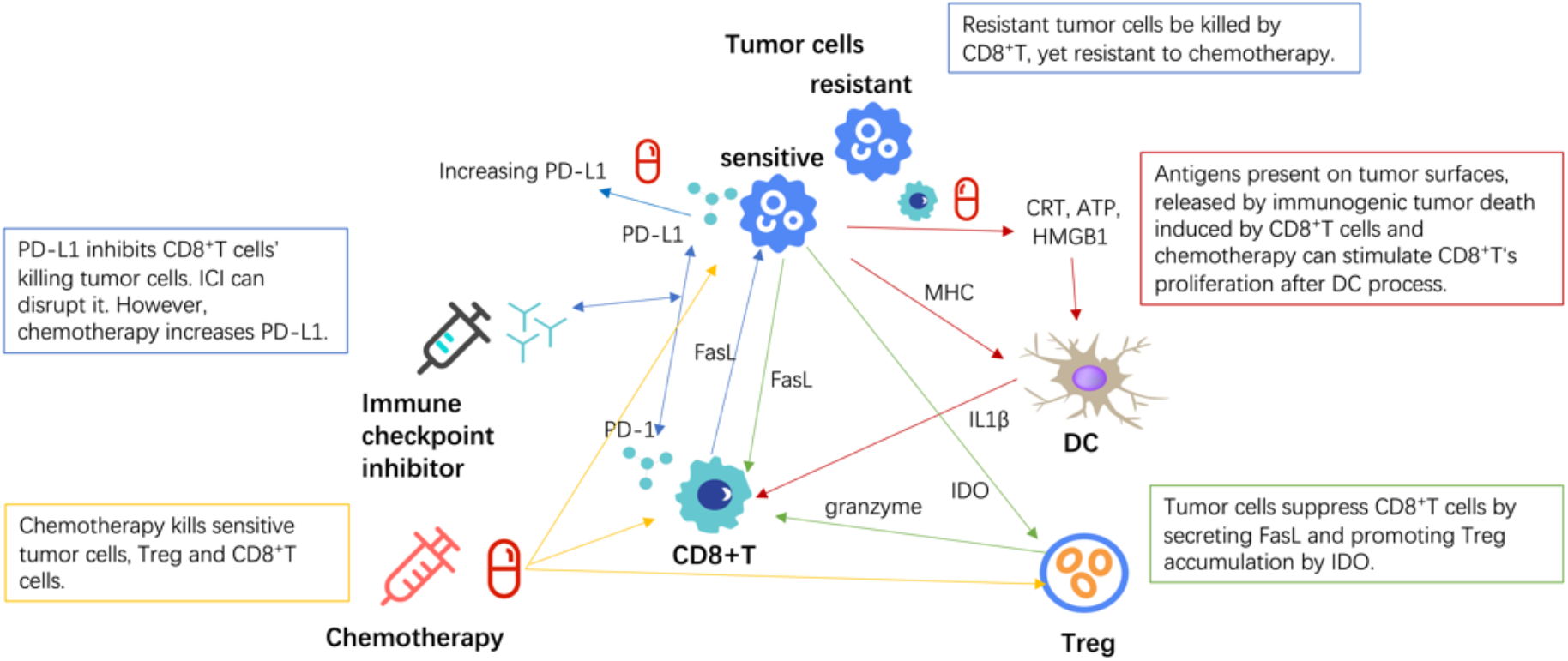
Overview of the ODE-based TIME dynamic evolutionary model. This chart shows the interplay of tumor cells, immune cells, and drugs.

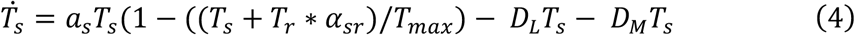

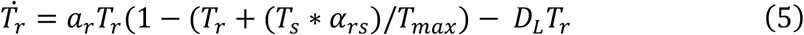

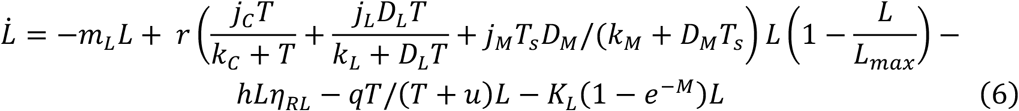

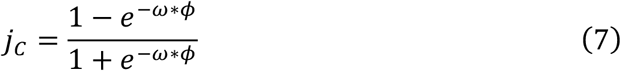

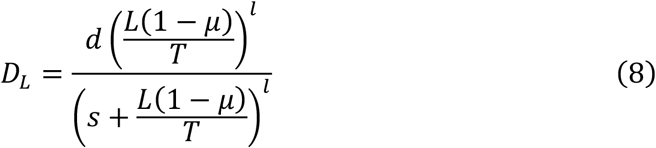

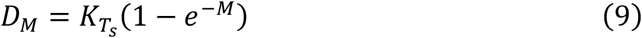

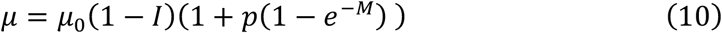

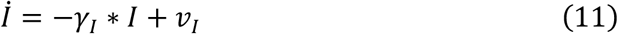

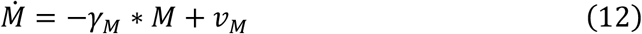

### Data

We collect two datasets of melanoma: The TCGA SKCM dataset (n=473) and the dataset published by Liu (n=120).^31,33^ Both datasets comprise the patient’s gene expression profiles, from which we can characterize the patient’s TIME in our ODE model (Table 1; refer to supplementary material). Patients in the TCGA SKCM dataset were treated with radiotherapy or chemotherapy. Patients in the Liu dataset were treated with ICIs. We use these two datasets to fit the parameters characterizing the TIME with and without treatment.

**Table 1.** Biomarkers, computational immunogenomic approaches, and observable variables.

| **Biomarkers** | **PD-1 expression** | **Antigen presentation molecules expression** | **Regulatory T cells to CD8<sup>+</sup>T cells ratio** | **CD8<sup>+</sup>T cell signatures expression** |
| --- | --- | --- | --- | --- |
| **Computational immunogenomic approaches** | Associated genes expression | MHC-I and MHC-II associated HLA genes ssGSEA | TIME composition by CIBERSORT | Single cell analysis |
| **Observable variables in our ODE model** | Fraction of CD8<sup>+</sup>T cells suppressed by ICs ($\mu_0$) | Normalized ssGSEA score of antigen presentation on the tumor surface ($\phi$) | Ratio of regulatory T cells to CD8<sup>+</sup>T cells ($\eta_{RL}$) | Ratio of CD8<sup>+</sup>T cells to tumor cells ($\eta_{LT}$) |

### Parameter estimation

Viewing our ODE-based TIME dynamic evolutionary model as a function, *F. y*_*it*_ *= F*(*x*_*i*_, *y*_*i*0_, *θ, z*_*i*_). *y*_*i*0_ and *y*_*it*_ are the TIME state value of a patient *i* at initial time and t, respectively. *x*_*i*_ are observable patient-specific variables, *z*_*i*_ are unobservable variables, and *θ* are parameters. We aim to estimate *θ, z*_*i*_ by combining data including *x*_*i*_ and *y*_*it*_.

We use the TCGA SKCM dataset to fit the TIME parameters without treatment. We choose the ratio of CD8^+^T cell population to tumor cell population (*η*_*LT*_) at first diagnosis as the target variable to be predicted. The initial value of *T*_*s*_, *T*_*r*_ and *L* are set to 1 × 10^4^, 4 × 10^1^, and 1 × 10^2^ respectively. Following the literature, we assume the intrinsic growth rates of sensitive and resistant cancer cells as *a*_*s*_ and *a*_*r*_, where *a*_*s*_ < *a*_*r*_. The simulated CD8^+^T cell population (*L*_*t*_) is determined by the ODE model characterized by *x*_*i*_, *z*_*i*_ and *θ*.

The presence and processing of antigens increase the growth rate of CD8^+^T cells, while the populations of tumor cells and regulatory T cells decrease the growth rate of CD8+T cells. At the early stage of tumor growth, the primary tumor antigens that stimulate T cell response are tumor antigens expressed on tumor cells.^5^ The proliferation rate of CD8^+^T cells is *rj*_*C*_ *T/*(*k*_*C*_ + *T*), where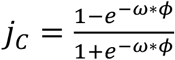. The regulatory effects from tumor cells and regulatory T cells are represented as *q* and *hη*_*RL*_. Consequently, the observable patient-specific variables (*x*_*i*_) include the expression of MHC-I and MHC-II molecules (measured by the ssGSEA score *ϕ*, a normalized summation of both molecules) and the ratio of regulatory T cells to CD8^+^T cells (*η*_*RL*_). The unobservable variable (*z*_*i*_) is the patient-specific antigens process ability (*r*). The parameters to be estimated from data include the regulatory effects on CD8+T cells by tumor cells (*q*) and regulatory T cells (*h*) and a scaling factor *ω*.

We use the Liu dataset to fit the TIME parameters with ICI treatment. The response result (response or disease progression) of ICI treatment is the target variable to be predicted. The observable patient-specific variables *x*_*i*_ include the fraction of CD8^+^T cells suppressed by checkpoints (µ_0_), which is measured by the PD-1 expression rate of the tumor. The fractional cell kill coefficient of CD8^+^T cells on tumor cells (*d*) and the rate of antigen presence by CD8^+^T cells induced immunogenetic tumor death (*j*_*L*_) are two non-patient-specific parameters affecting the efficacies of ICI treatment. *d* is adopted from the literature. *j*_*L*_ is fitted by the Liu dataset.

There are two critical parameters related to the chemotherapy: the rate of antigen presence by chemotherapy-induced immunogenetic tumor death (*j*_*M*_) and upregulation rate of chemotherapy drugs on checkpoint inhibitors (*p*). *j*_*M*_ stimulates the proliferation of CD8^+^T cells. *p* determines how the use of chemotherapy drugs increases the expression of checkpoint inhibitors. Please refer to the supplementary material for details of the parameter setting.

### Chemotherapy and ICI combination decision

The agent makes decisions based on the patient’s TIME state *S*_*t*_, derived from our ODE model. The following describes the RL algorithm’s decision epochs, action space, and reward function.

### Decision epochs *T*_*epoch*_

Following clinical trial KEYNOTE-189, patients are assigned to receive platinum-based drug (chemotherapy) plus pembrolizumab (PD-1 Inhibitor) or placebo every three weeks for four cycles, followed by up to 35 cycles of pembrolizumab or placebo.^45^ Consequently, we formed a decision process with four decision epochs at a three-week interval. After the first four cycles, the agent will repeat the last ICI decision for up to six cycles if a complete response (CR) is not achieved. And if the tumor cells’ population is <1 × 10^2^(CR) or the tumor cells’ population is >2.6 × 10^8^(number of cells for an 8mm diameter ball shape tumor) before four cycles (treatment failure), the treatment will be ended, thus *T*_*epoch*_ *≤* 4.

### Action space *A*

At each decision epoch, the doctor decides (a) whether or not to use the ICI and (b) the chemotherapy drug doses. The maximum dosage of the chemotherapy drug is set to be five, indicating that the chemotherapy drug is fully saturated. The saturation term (1 *− e*^*™M*^) reaches one when M equals five.

### Reward r

The decision process rewards the tumor size reduction and least ICI usage for economic cost consideration. Specifically, our reward function comprises immediate reward *r*_*t*_ and terminal reward *r*_*T*_. Immediate reward *r*_*t*_ is tumor size reduction rate.

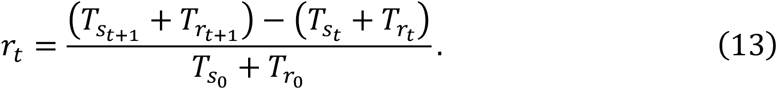

As for the terminal reward, if the final tumor cells’ population is lower than 1 × 10^2^ (tumor elimination), the terminal reward is maxed at 1. In the case of tumor elimination, the cost of ICI *I*_*t*_ will not be calculated. Otherwise, if the tumor cells’ population is more extensive than 2.6 × 10^8^ (treatment failure), the terminal reward is 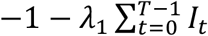, where *λ*_1_ is the cost for ICI. The additional term *λ*_2_(4 *− T*) rewards the early tumor elimination before the fourth cycle. If the final tumor cells’ population is between 1 × 10^2^and 2.6 × 10^8^, we will simulate the TIME without treatment for another six cycles (18 weeks) at *T*^’^ (4 < *T*^’^ *≤* 10) and calculate the tumor’s re-growth rate. This additional simulation characterizes the delayed response of ICI and the evolutionary dynamics of resistant and sensitive cancer cells.^34^

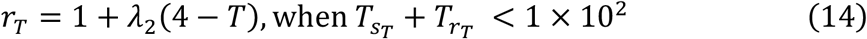

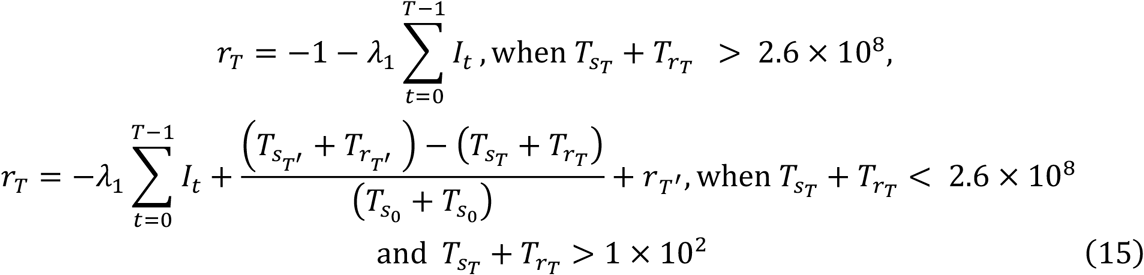

### Reinforcement Learning training

We train a deep reinforcement learning (DRL) agent by deep deterministic policy gradient (DDPG) to interact with simulated TIME.^46^ There are two main reasons for using this algorithm. First, in our decision optimization task, the dose of the chemotherapy drug is continuous, so we need an algorithm capable of operating continuous action space. Second, the TIME simulation is computationally expensive and slow, so an off-policy algorithm is preferred. We train the agent for 10000 steps. To create a diversity of simulated TIME, we train 200 TIME simulators corresponding to 200 patients in the TCGA SKCM cohort. Then, we use another cohort of TCGA SKCM patient simulators (n=30) to evaluate the performance of the DRL-derived policy.

## RESULTS

### Evolutionary dynamics of TIME

The real data fits our ODE model well. Without ICI treatment, the mean absolute errors between the ratio of CD8^+^T cells to tumor cells calculated by our model and that estimated by the CD8^+^T cell signatures expression are 0.003 and 0.0015 for the TCGA SKCM dataset and Liu dataset, respectively. With ICI treatment, our model can predict the response of ICI therapy with an accuracy of 0.63 and a recall of 0.64 in the Liu dataset. These results indicate that the proposed ODE model can well characterize the evolutionary dynamics of the TIME with and without treatment of ICI.

The fitted parameters have biological insights into the various mechanisms of CD8^+^T cells’ proliferation. The average CD8^+^T cells proliferation rate stimulated by antigens presence on tumor surface is 0.1 (*rj*_*C*_). Considering the natural CD8^+^T cells death rate *m*_*L*_, 0.05, if there does not exist suppression from tumor cells and regulatory T cells, the CD8^+^T cells will increase at the average rate, 0.05, higher than the growth rates of sensitive and resistant tumor cells. However, the suppressive effect from tumor cells (*q*) and regulatory T cells (*h*) exist, and the former one is three times the latter one (0.03 vs. 0.01). This indicates that chemotherapy can promote the CD8+T cell proliferation rate by reducing the suppression of tumor cells. The average CD8^+^T cells proliferation rate stimulated by CD8^+^T cells induced tumor immunogenetic death (*rj*_*L*_) is 0.016. This rate is much lower than *rj*_*C*_ (0.1), indicating that the CD8^+^T cell proliferation is primarily stimulated by antigens on the tumor surface. If the tumor is too “cold,” with extremely low levels of tumor surface antigens, removing tumor cell suppression and CD8+T cell stimulation from tumor immunogenetic death may not help increase the infiltration of CD8+T cell.

Given TCGA SKCM patients’ specific biomarkers, the ODE model can estimate the tumor growth and TIME evolutionary process before diagnosis and anticipate the subsequent evolutionary dynamics with treatment intervention. From our ODE model’s dynamics, we found that characterizing patients’ TIME heavily depends on the ratio of CD8^+^T cell population to tumor cell population (*η*_*LT*_) and CD8^+^T cell growth rate. Based on these factors, we can classify the TIME of patients into three categories: “hot tumor”, “cold tumor”, and “extremely cold tumor”.^5^ For hot tumor, there exists sufficient tumor surface antigen (high MHC ssGSEA score *ϕ*), resulting in high CD8^+^T infiltration *η*_*LT*_ (high expression of CD8+T cells signatures). Cold tumor is insufficiently inflamed, yet it can be turned hot. The extremely cold tumor has an extremely low level of tumor surface antigen and CD8^+^T cell infiltration, with little chance of being turned hot. In the following, we use three representative patients to illustrate the differences in these three categories.

Patients TCGA-DA-A1IB, TCGA-EB-A4XL, and TCGA-EE-A2GR are examples of hot, cold, and extremely cold tumors, respectively. In Figure 2, the green points represent the CD8^+^T cell population at diagnosis, estimated by the expression of the CD8+T cell signatures. In contrast, the green curves represent the CD8^+^T cell population estimated by our model. To summarize, we find that the CD8+T cell population grows quite fast in hot tumors (Figure 2a); CD8+T cells grow faster than tumor cells at the beginning and later, similar to tumor cells in cold tumors (Figure 2b); CD8+T cells grow slower than tumor cells in the extremely cold tumor (Figure 2c). These differences are reflected by the ratio of the CD8^+^T cell population to tumor cell population curves with different slopes (Figure 2d).

**Figure 2.**
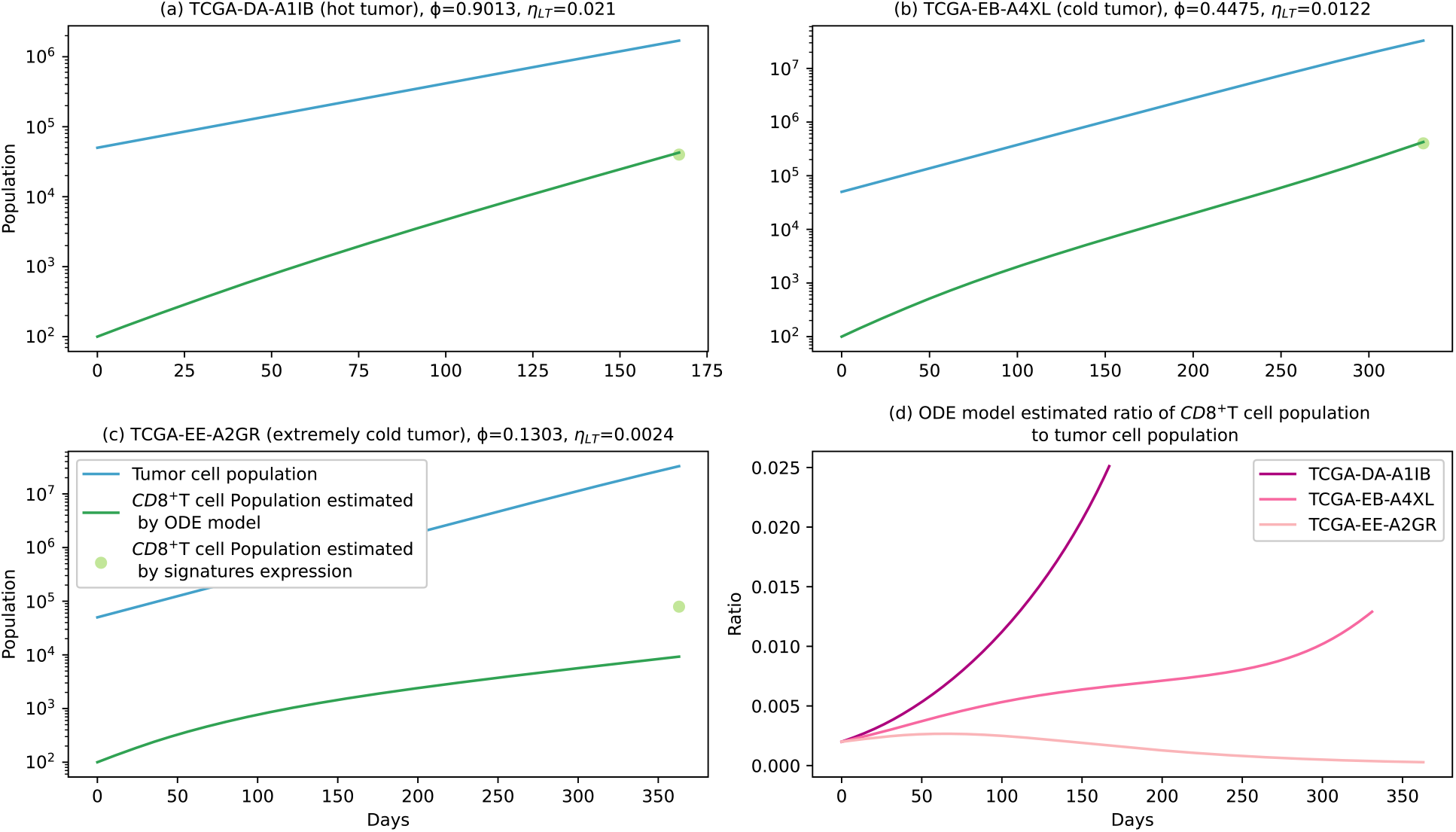
Simulated TIME dynamics before the diagnosis of representative TCGA SKCM patients: (a) TCGA-DA-A1IB (Hot tumor), (b) TCGA-EB-A4XL (cold tumor), (c) TCGA-EE-A2GR (extremely cold tumor). (d) The ODE model estimated the ratio of the CD8+T cell population to the tumor cell population.

### Comparison of fixed ICC schedules and DRL-derived schedules

This section compared different fixed ICC schedules on 30 TCGA SKCM patients in the test set (Figure 3). For instance, (1,2) means that 1 unit of ICI combined with two units of chemotherapy drug is administrated for four cycles, followed by six cycles of ICI. With ICI, the average reward increases when chemotherapy is at 1, 2, or 3 units (0.922 vs. 0.657, 0.875 vs. 0.856, 0.824 vs. 0.712). However, the average reward decreases when chemotherapy is 4 or 5 (0.595 vs. 0.692, 0.399 vs. 0.596). These findings indicate that high dosages of chemotherapy hurt anti-tumor immunity, leading to the inefficacy of ICI. The average reward peaks when the chemotherapy dosage is one unit with ICI (0.922). Such a tradeoff does not apply to the extremely cold tumor case in Figure 3, where combining ICI with chemotherapy leads to worse rewards. Instead, a high dosage of chemotherapy alone can decrease sensitive tumor cells at its best. Notably, our DRL agent yields the highest overall average reward (1.079).

**Figure 3.**
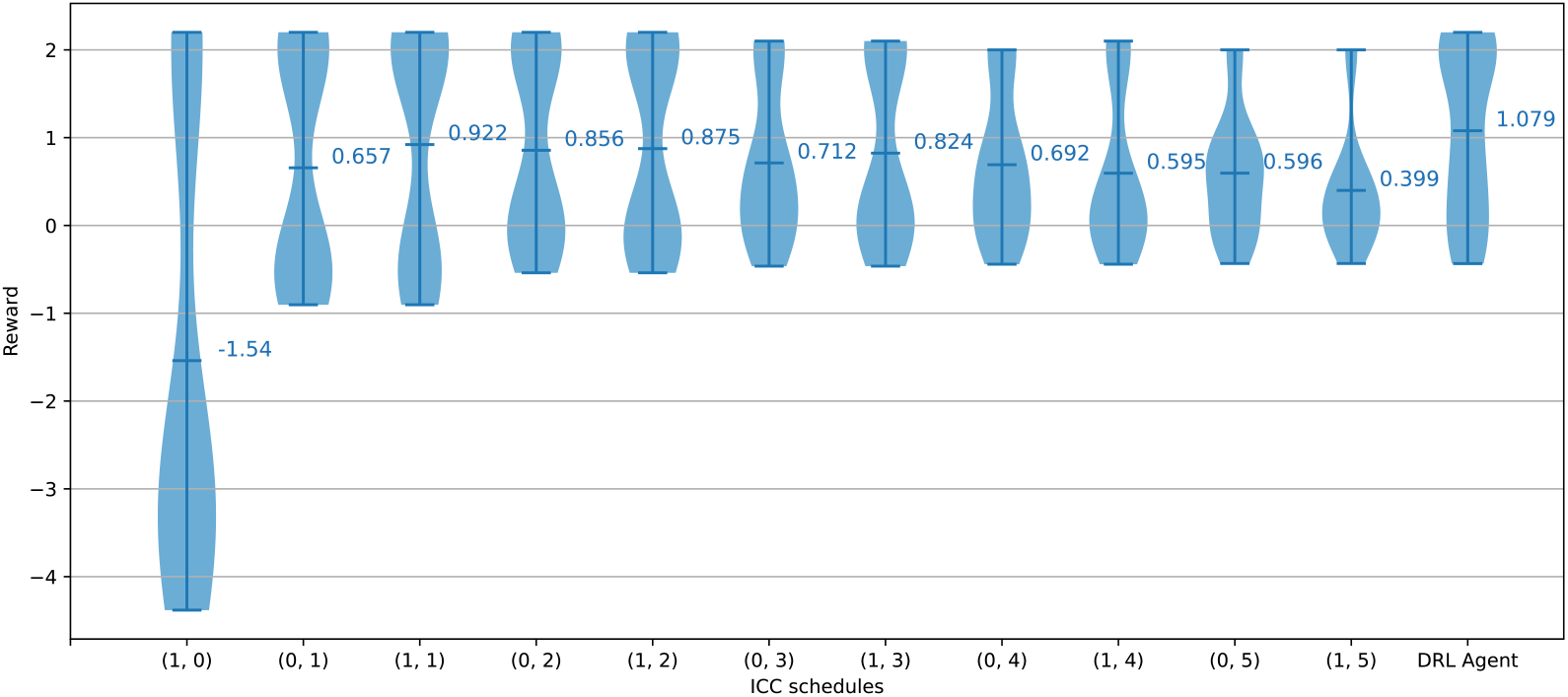
Performance of fixed ICC schedules and DRL-derived ICC schedules. For the ICC schedules, (*a, b*) indicates *a* unit of ICI combined with *b* units of chemotherapy drug are administrated. The violin plot shows the distribution and mean value of simulated ICC reward on 30 TCGA SKCM patients in the test set.

Results reported in Figure 3 indicate that fixed ICC schedules cannot handle the complex tradeoff between reducing tumor size and enhancing the CD8^+^T cell population, which is highly personalized. Our proposed DRL agent can learn the optimal schedule at specific TIME state *S*_*t*_ for individual patients. To summarize, we find that its decision rules are mainly associated with sensitive tumor cell population, resistant tumor cell population, and the ratio of CD8^+^T cell population to tumor cell population (*η*_*LT*_), represented by the three dimensions in Figure 4(a). In Figure 4 (a), the color palette and green surface indicate the DRL-derived dosage of chemotherapy and the ICI usage decision boundary, respectively. When the CD8^+^T cell infiltration (*η*_*LT*_) increases (“hotter” tumor), a lower dosage of chemotherapy combined with ICI is suggested. Another pattern is that increasing the population of resistant tumor cells makes the ICI suggested in lower CD8^+^T cell infiltration (*η*_*LT*_) area. To demonstrate the 4-cycle sequential treatment decisions of TCGA SKCM patients in the test set in the 2-dimensional figure, we fix the resistant tumor cell population to the 2.98 × 10^3^ (the average of all patients) and visualize the dimension of sensitive tumor cell population and the ratio of CD8^+^T cell population to tumor cell population (*η*_*LT*_) in Figure 4 (b). Similarly, the background color indicates the agent-derived dosage of chemotherapy. Lines with arrows indicate TCGA SKCM patients’ TIME dynamics.

**Figure 4.**
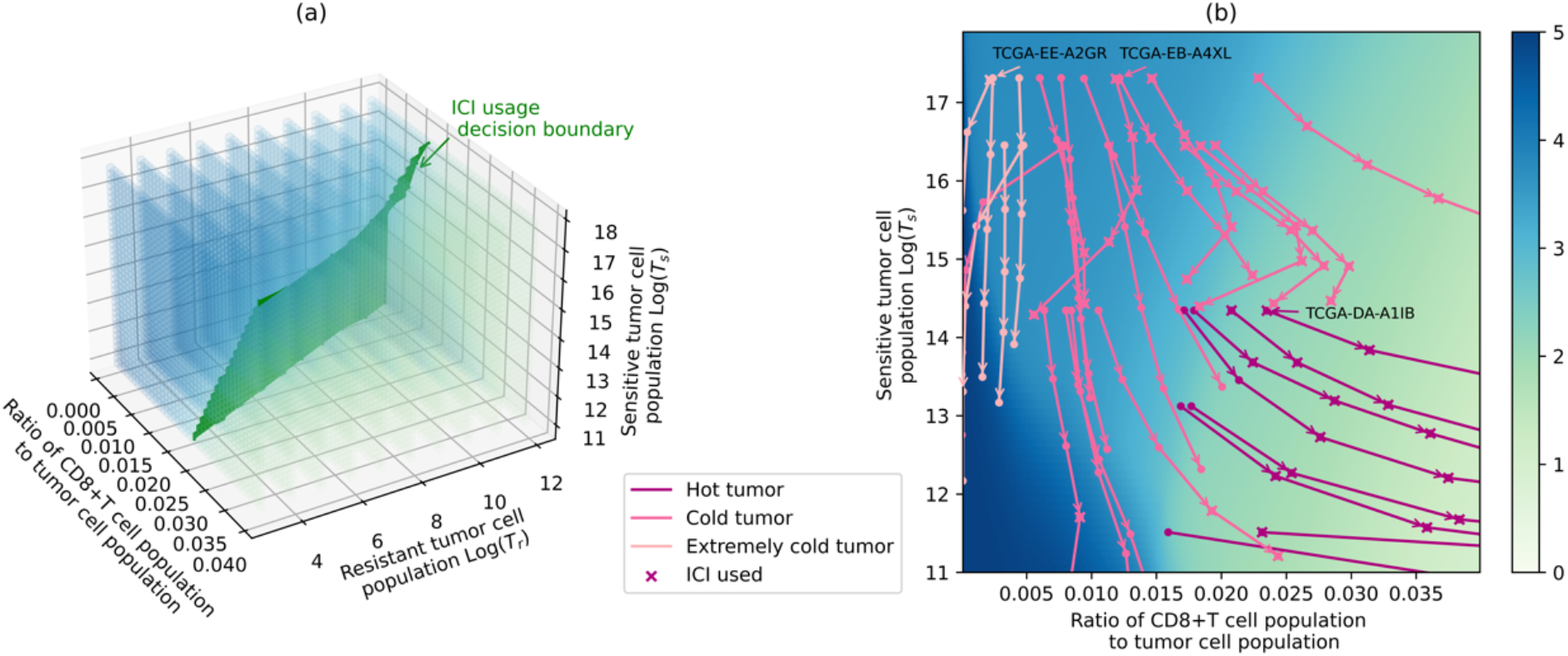
Visualization of the DRL-derived ICC schedules. (a) Three-dimensional visualization of the DRL-derived chemotherapy dosage and ICI usage decision boundary. The color palette (from blue to light green) indicates the DRL-derived dosage of chemotherapy (from high to low). The green surface is the ICI usage decision boundary (not use vs. use, left vs. right). (b) Two-dimensional visualization of DRL-agent-derived ICC schedules and TCGA SKCM patients’ TIME dynamics with four cycles of ICC intervention. The plane color indicates the DRL-derived dosage of chemotherapy. The TIME dynamics are indicated by arrows, for which the color tells the category the patient’s TIME belongs to, and the marker “X” shows the ICI used at the corresponding cycle.

Specifically, patients with extremely cold tumors (pink) are initially located in low CD8^+^T cell infiltration (*η*_*LT*_) area, where a high dosage of chemotherapy is suggested. For these patients, a high dosage of chemotherapy can reduce the tumor size, and the CD8^+^T cell infiltration (*η*_*LT*_) remains low. As demonstrated by the downward pink arrows, sensitive tumor cells are being killed by applying chemotherapy.

Conversely, patients with hot tumors (rose) are initially located on the right side of ICI decision boundaries, where ICI combined with a low dosage of chemotherapy is suggested. This ICC combination strategy stimulates CD8^+^T cells’ proliferation and significantly reduces the tumor size, as the rose arrows pointing to the lower right demonstrate.

Patients with cold tumors (dark pink) are initially located at the left side of decision boundaries, and the DRL agent suggests a middle-level dosage of chemotherapy. When the TIME is turned “hot” enough (by the middle-level chemotherapy) to be favorable for ICI treatment, ICC with a low dosage of chemotherapy will be applied. However, not all cold tumors can be turned “hot” enough for the following ICC. Seven patients are suggested single chemotherapy, as the downward dark pink arrows demonstrate. Three patients benefited from ICC consistently, as shown by the dark pink arrows pointing to the lower right. Seven patients have failed the ICC due to the high resistant tumor cell growth rate, which decreases CD8^+^T infiltration (*η*_*LT*_). This is demonstrated by the dark pink arrows moving to the left at the third or fourth cycle.

To examine the TIME dynamics of different types of patients in more detail, we present the case of four representative patients in Figure 5. TCGA-EE-A2GR is an example of an extremely cold tumor, and his/her suggested treatment is a high dosage of chemotherapy drug without ICI. TCGA-DA-A1IB is an example of a hot tumor.

**Figure 5.**
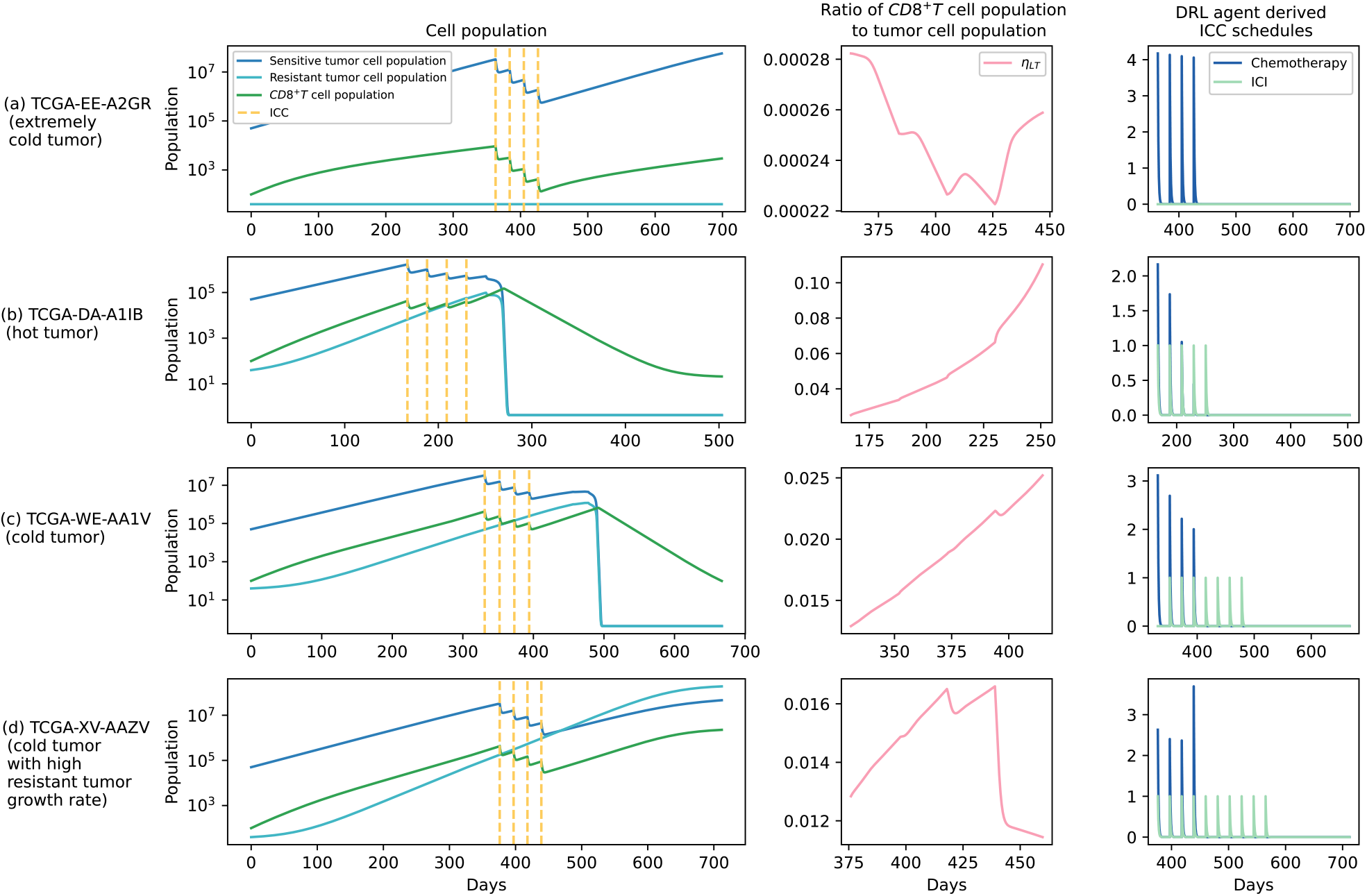
TIME evolutionary dynamics simulation with DRL-derived ICC schedules, ratio of CD8^+^T cell population to tumor cell population, and DRL-derived ICC schedules for representative patients: (a) TCGA-EE-A2GR (extremely cold tumor); (b) TCGA-DA-A1IB (hot tumor); (c) TCGA-EB-A4XL (cold tumor); (d) TCGA-XV-AAZV (cold tumor with high resistant tumor growth rate).

Two cycles of low-dosage chemotherapy effectively help stimulate the CD8^+^T cell proliferation. Then, the ICI wipes out the tumor cells in the following cycles. TCGA-EB-A4XL is an example of a cold tumor that can be turned hot. With one cycle of middle-dosage chemotherapy, the CD8^+^T infiltration (*η*_*LT*_) increases to enable the following ICC with a lower dosage of chemotherapy. TCGA-XV-AAZV is an example of a cold tumor that cannot be turned hot. This patient’s tumor has a high resistant tumor growth rate. The rapidly increasing proportion of resistant tumor cells decreases the ratio of the CD8+T cell population to the tumor cell population, leading to the failure of ICI.

## DISCUSSION

The current study sheds light on how to leverage real-world clinical data and DRL to develop personalized optimal ICC by characterizing the complex biological dynamics of the patient’s TIME. Our ODE-based TIME dynamic model provides a theoretical framework for exploring the best use of ICI. The DRL model proposed here has the potential to guide the personalized ICC schedules to steer dynamic TIME evolution and achieve the optimal treatment outcome for individual patients.

Our study attempts to address the dilemma clinicians face while deciding to use ICC for individual patients. (a) Whether the chemotherapy should be administrated at an MTD? The dosage of chemotherapy depends on the individual patient’s TIME. High dosage is only suggested for patients with extremely low CD8^+^T cell infiltration (extremely cold tumor). For cold and hot tumors, an appropriate dosage of chemotherapy can help induce CD8+T cell proliferation. (b) Whether a patient may have a response to ICI? Our model suggests prior patient stratification based on their TIME. For patients with extremely cold tumors or cold tumors with fast resistant cell growth, the combination with ICI probably does not help. (c) In what sequence should ICI and chemotherapy be administrated? The ICI usage favors “hot” TIME, that is, tumors with high CD8^+^T cell infiltration. Consequently, for a hot tumor, chemotherapy and ICI are suggested simultaneously. In contrast, for a cold tumor, the middle dosage of chemotherapy is suggested at first several cycles to turn the TIME “hotter,” and then ICI is suggested to be combined. In short, CD8^+^T cell infiltration and tumor resistance to chemotherapy are critical factors for ICC schedule design. We recommend that healthcare practitioners acquire associated information from CD8^+^T cell signatures expression and in vitro drug sensitivity and resistance test (DSRT)^47^ while determining the ICC schedule to maximize the likelihood of treatment outcome.

Our study has limitations. First, although we used the real-world genomic and transcriptomic features to calibrate the model, our model is still largely theoretical. We call for more in vitro and in vivo experiments and clinical trials to verify and extend this theoretical model. Second, due to a lack of information in the data, we did not consider the variety of chemotherapy drugs in the model. Future research is needed to examine the different combination strategies with varying drugs of chemotherapy. Third, our study was focused on melanoma. The utility of our model in other cancer types needs relevant data to validate.

## Supporting information

Supplementary ODE model parameters setting, Supplementary reward shaping for ICC decision

## Data Availability

All data are public. Raw code is available at GitHub.

https://github.com/yaoyao0111/ICC_decision.git

## Data and code availability statement

All data are public. Raw code is available at https://github.com/yaoyao0111/ICC_decision.git

## Ethical statement

This research has been approved by the research ethics committee of CityU (HU-STA-00000380).

## REFERENCES

1. Topalian SL, Drake CG, Pardoll DM. Immune checkpoint blockade: a common denominator approach to cancer therapy. Cancer cell. 2015;27(4):450–61.

2. Postow MA, Callahan MK, Wolchok JD. Immune checkpoint blockade in cancer therapy. Journal of clinical oncology. 2015;33(17):1974.

3. Webster RM. The immune checkpoint inhibitors: where are we now? Nature reviews Drug discovery. 2014;13(12):883.

4. Zhang Y, Chen L. Classification of advanced human cancers based on tumor immunity in the microenvironment (TIME) for cancer immunotherapy. JAMA oncology. 2016;2(11):1403–4.

5. Bonaventura P, Shekarian T, Alcazer V, Valladeau-Guilemond J, Valsesia-Wittmann S, Amigorena S, et al. Cold tumors: a therapeutic challenge for immunotherapy. Frontiers in immunology. 2019;10:168.

6. Galluzzi L, Humeau J, Buqué A, Zitvogel L, Kroemer G. Immunostimulation with chemotherapy in the era of immune checkpoint inhibitors. Nature reviews Clinical oncology. 2020;17(12):725–41.

7. Heinhuis KM, Ros W, Kok M, Steeghs N, Beijnen JH, Schellens JHM. Enhancing antitumor response by combining immune checkpoint inhibitors with chemotherapy in solid tumors. Annals of Oncology. 2019;30(2):219–35.

8. Leonetti A, Wever B, Mazzaschi G, Assaraf YG, Rolfo C, Quaini F, et al. Molecular basis and rationale for combining immune checkpoint inhibitors with chemotherapy in non-small cell lung cancer. Drug Resistance Updates. 2019;46:100644.

9. Salas-Benito D, Pérez-Gracia JL, Ponz-Sarvisé M, Rodriguez-Ruiz ME, Martínez-Forero I, Castañón E, et al. Paradigms on immunotherapy combinations with chemotherapy. Cancer discovery. 2021;11(6):1353–67.

10. Casares N, Pequignot MO, Tesniere A, Ghiringhelli F, Roux S, Chaput N, et al. Caspase-dependent immunogenicity of doxorubicin-induced tumor cell death. The Journal of experimental medicine. 2005;202(12):1691–701.

11. Barbon CM, Yang M, Wands GD, Ramesh R, Slusher BS, Hedley ML, et al. Consecutive low doses of cyclophosphamide preferentially target Tregs and potentiate T cell responses induced by DNA PLG microparticle immunization. Cellular immunology. 2010;262(2):150–61.

12. Vincent J, Mignot G, Chalmin F, Ladoire S, Bruchard M, Chevriaux A, et al. 5-Fluorouracil selectively kills tumor-associated myeloid-derived suppressor cells resulting in enhanced T cell–dependent antitumor immunity. Cancer research. 2010;70(8):3052–61.

13. Schiavoni G, Sistigu A, Valentini M, Mattei F, Sestili P, Spadaro F, et al. Cyclophosphamide synergizes with type I interferons through systemic dendritic cell reactivation and induction of immunogenic tumor apoptosis. Cancer research. 2011;71(3):768–78.

14. Tanaka H, Matsushima H, Nishibu A, Clausen BE, Takashima A. Dual therapeutic efficacy of vinblastine as a unique chemotherapeutic agent capable of inducing dendritic cell maturation. Cancer research. 2009;69(17):6987–94.

15. Cortes J, Cescon DW, Rugo HS, Nowecki Z, Im SA, Yusof MM, et al. KEYNOTE-355: Randomized, double-blind, phase III study of pembrolizumab+ chemotherapy versus placebo+ chemotherapy for previously untreated locally recurrent inoperable or metastatic triple-negative breast cancer. American Society of Clinical Oncology; 2020.

16. Paz-Ares L, Vicente D, Tafreshi A, Robinson A, Parra HS, Mazières J, et al. A randomized, placebo-controlled trial of pembrolizumab plus chemotherapy in patients with metastatic squamous NSCLC: protocol-specified final analysis of KEYNOTE-407. Journal of Thoracic Oncology. 2020;15(10):1657–69.

17. Gadgeel S, Rodríguez-Abreu D, Speranza G, Esteban E, Felip E, Dómine M, et al. Updated analysis from KEYNOTE-189: pembrolizumab or placebo plus pemetrexed and platinum for previously untreated metastatic nonsquamous non-small-cell lung cancer. Journal of clinical oncology. 2020;38(14):1505–17.

18. Crawford J, Dale DC, Lyman GH. Chemotherapy-induced neutropenia: risks, consequences, and new directions for its management. Cancer. 2004;100(2):228– 37.

19. Rosenblatt M. The large pharmaceutical company perspective. New England Journal of Medicine. 2017;376(1):52–60.

20. Xu Y, Hosny A, Zeleznik R, Parmar C, Coroller T, Franco I, et al. Deep learning predicts lung cancer treatment response from serial medical imaging. Clinical Cancer Research. 2019;25(11):3266–75.

21. Yang J, Li Z, Wu WKK, Yu S, Xu Z, Chu Q, et al. Deep learning identifies explainable reasoning paths of mechanism of action for drug repurposing from multilayer biological network. Briefings in Bioinformatics. 2022;23(6):bbac469.

22. de Pillis LG, Gu W, Radunskaya AE. Mixed immunotherapy and chemotherapy of tumors: modeling, applications and biological interpretations. Journal of theoretical biology. 2006;238(4):841–62.

23. Yin A, Moes DJAR, Hasselt JGC, Swen JJ, Guchelaar H. A Review of Mathematical Models for Tumor Dynamics and Treatment Resistance Evolution of Solid Tumors. CPT Pharmacometrics Syst Pharmacol. 2019 Oct;8(10):720–37.

24. Jarrett AM, Faghihi D, Hormuth DA, Lima EA, Virostko J, Biros G, et al. Optimal control theory for personalized therapeutic regimens in oncology: Background, history, challenges, and opportunities. Journal of clinical medicine. 2020;9(5):1314.

25. Khalili P, Vatankhah R. Optimal control design for drug delivery of immunotherapy in chemoimmunotherapy treatment. Computer Methods and Programs in Biomedicine. 2023;229:107248.

26. Padmanabhan R, Meskin N, Haddad WM. Reinforcement learning-based control of drug dosing for cancer chemotherapy treatment. Mathematical biosciences. 2017;293:11–20.

27. Siewe N, Friedman A. Combination therapy for mCRPC with immune checkpoint inhibitors, ADT and vaccine: A mathematical model. PLoS One. 2022;17(1):e0262453.

28. Butner JD, Wang Z, Elganainy D, Al Feghali KA, Plodinec M, Calin GA, et al. A mathematical model for the quantification of a patient’s sensitivity to checkpoint inhibitors and long-term tumour burden. Nature biomedical engineering. 2021;5(4):297–308.

29. Kim Y, Choe BY, Suh TS, Sung W. A Mathematical Model for Predicting Patient Responses to Combined Radiotherapy with CTLA-4 Immune Checkpoint Inhibitors. Cells. 2023;12(9):1305.

30. Butner JD, Elganainy D, Wang CX, Wang Z, Chen SH, Esnaola NF, et al. Mathematical prediction of clinical outcomes in advanced cancer patients treated with checkpoint inhibitor immunotherapy. Science advances. 2020;6(18):eaay6298.

31. Liu D, Schilling B, Liu D, Sucker A, Livingstone E, Jerby-Arnon L, et al. Integrative molecular and clinical modeling of clinical outcomes to PD1 blockade in patients with metastatic melanoma. Nature medicine. 2019;25(12):1916–27.

32. Eastman B, Przedborski M, Kohandel M. Reinforcement learning derived chemotherapeutic schedules for robust patient-specific therapy. Scientific Reports. 2021;11(1):17882.

33. Tomczak K, Czerwińska P, Wiznerowicz M. Review The Cancer Genome Atlas (TCGA): an immeasurable source of knowledge. Contemporary Oncology/Współczesna Onkologia. 2015;2015(1):68–77.

34. Zhang J, Cunningham JJ, Brown JS, Gatenby RA. Integrating evolutionary dynamics into treatment of metastatic castrate-resistant prostate cancer. Nature communications. 2017;8(1):1816.

35. Wainwright DA, Balyasnikova IV, Chang AL, Ahmed AU, Moon KS, Auffinger B, et al. IDO Expression in Brain Tumors Increases the Recruitment of Regulatory T Cells and Negatively Impacts SurvivalIDO Regulates Treg Infiltration in Brain Tumors. Clinical cancer research. 2012;18(22):6110–21.

36. Choi BD, Gedeon PC, Herndon JE, Archer GE, Reap EA, Sanchez-Perez L, et al. Human Regulatory T Cells Kill Tumor Cells through Granzyme-Dependent Cytotoxicity upon Retargeting with a Bispecific AntibodyRedirected Regulatory T Cells Kill Tumors. Cancer immunology research. 2013;1(3):163–7.

37. Liu YT, Sun ZJ. Turning cold tumors into hot tumors by improving T-cell infiltration. Theranostics. 2021;11(11):5365.

38. Principe DR, Chiec L, Mohindra NA, Munshi HG. Regulatory T-cells as an emerging barrier to immune checkpoint inhibition in lung cancer. Frontiers in oncology. 2021;11:684098.

39. Chen B, Khodadoust MS, Liu CL, Newman AM, Alizadeh AA. Profiling tumor infiltrating immune cells with CIBERSORT. Cancer Systems Biology: Methods and Protocols. 2018;243–59.

40. Gajewski TF, Schreiber H, Fu YX. Innate and adaptive immune cells in the tumor microenvironment. Nature immunology. 2013;14(10):1014–22.

41. Wang Q, Ju X, Wang J, Fan Y, Ren M, Zhang H. Immunogenic cell death in anticancer chemotherapy and its impact on clinical studies. Cancer letters. 2018;438:17–23.

42. Jaime-Sanchez P, Uranga-Murillo I, Aguilo N, Khouili SC, Arias MA, Sancho D, et al. Cell death induced by cytotoxic CD8+ T cells is immunogenic and primes caspase-3–dependent spread immunity against endogenous tumor antigens. Journal for immunotherapy of cancer. 2020;8(1).

43. Srinivasan B. A guide to the Michaelis–Menten equation: steady state and beyond. The FEBS journal. 2022;289(20):6086–98.

44. Bailly C, Thuru X, Quesnel B. Combined cytotoxic chemotherapy and immunotherapy of cancer: modern times. NAR cancer. 2020;2(1):zcaa002.

45. Gandhi L, Rodríguez-Abreu D, Gadgeel S, Esteban E, Felip E, De Angelis F, et al. Pembrolizumab plus chemotherapy in metastatic non–small-cell lung cancer. New England journal of medicine. 2018;378(22):2078–92.

46. Lillicrap TP, Hunt JJ, Pritzel A, Heess N, Erez T, Tassa Y, et al. Continuous control with deep reinforcement learning. arXiv preprint arXiv:150902971. 2015;

47. Popova AA, Levkin PA. Precision medicine in oncology: In vitro drug sensitivity and resistance test (DSRT) for selection of personalized anticancer therapy. Advanced Therapeutics. 2020;3(2):1900100.

