## Supplementary ODE model parameters setting, Supplementary reward shaping for ICC decision for "Optimized patient-specific immune checkpoint inhibitors therapy for cancer treatment based on tumor immune microenvironment modeling"

#### 1.1. Observations and experiments

Some parameters in our models can be estimated from clinical observations and experiments.

##### 1.1.1. Tumor intrinsic growth rate

Take melanoma as an example. At the T4 stage, the melanoma is thicker than 4mm. Our model assumes the largest tumor can grow to be an 8mm diameter ball-shaped bulk. It is commonly assumed that a cubed volume tumor contains $10^9$ cells, so we set the maximum population of tumor cells, $T_{max}$, as $2.6e^8$. The median and mean volume doubling time is 94/144 days for primary melanoma and 33/64 days for metastatic ones. For patients in the TCGA SKCM dataset, most of them have primary tumors, so we parametrize the sensitive tumor intrinsic growth rate $a_s$ ranging from $1.45e^{-2}$to $2.23e^{-2}$ so that the simulated sensitive tumor grows from 1mm to 2mm in around 94 to 144 days. Similarly, for patients in the Liu dataset, most tumors are metastatic. We assume this cohort of patients has tumors with a higher intrinsic growth rate, $a_s$, ranging from $2.23e^{-2}$ to $3.27e^{-2}$, so that the sensitive tumor grows from 1mm to 2mm in around 64 to 94 days.

Interaction between sensitive and resistant tumor cells was well modelled.[1] The competition coefficients in our model were directly adopted from this research, that is, $\alpha_{sr} = 0.7,\ \alpha_{rs} = 0.9$. In their model, the resistant tumor intrinsic growth rate is estimated to be twice the sensitive one. However, we assume that the ratio of resistant tumor intrinsic growth rate to sensitive ones ($\frac{a_r}{a_s}$) uniformly vary from 1.3 to 1.8, deriving the chemotherapy resistance heterogeneity. Here, we present the proportion of resistant tumor cell ($\eta_{rT}$) when the ratio of resistant tumor intrinsic growth rate to sensitive ones ($\frac{a_r}{a_s}$ ) is uniformly distributed over different intervals (Extended Figure 1). For instance, U (1.3,1.4) means that $\frac{a_r}{a_s}$ is uniformly distributed over [1.3, 1.4]. When $\frac{a_r}{a_s}$ distributed in intervals with higher value, the proportion of resistant tumor ($\eta_{rT}$) increases. According to Zhang et al., when the resistant tumor's proportion is larger than 20%, the patient will be non-responder to therapy and excluded from this therapy for ethical reasons.[2] Consequently, we set the uniform distribution interval as [1.3, 1.8], making most patient's proportion of resistant tumor is lower than 20%.

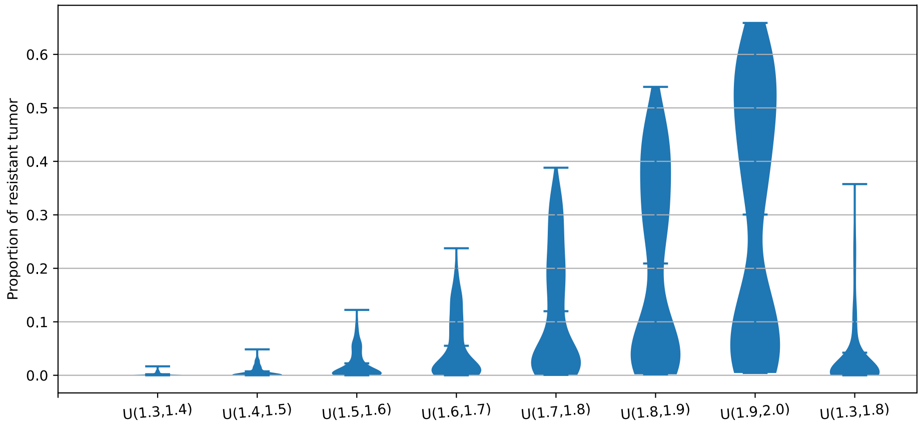


**Extended Figure 1.** Proportion of resistant tumor cell when ratio of resistant tumor intrinsic growth rate to sensitive ones is uniformly distributed over different intervals.

#### 1.1.2. Immune cells death rate

In vivo T cell dynamics have been recorded, and the CD8$^+$T cells death rate $m_L$ was estimated to be 0.05.[3]

### 1.2. Assumptions

Assumption 1: The maximum carry capacity of tumor-specific CD8$^+$T cells is $1e^7$.

Assumption 2: In our model, we adopt the Michaelis-Menten term to model some products' formation rate to the substrate concentration. Namely, the tumor's suppression effects on CD8$^+$T cells to the population of the tumor, the production rate of antigen to the population of the tumor, the CD8+T lysed tumor, and the cytotoxic drug killed tumor. When substrate concentration equals to the Michaelis constants, $u, k_1, k_2, k_3$, the formation rates reach half of their maximum value. Because there are no existing validated experiment results, we set $k_1, k_2, k_3$ to be $1e^4$ in our model. Because the suppressive inhibitors IDO occurs in tumors with CD8$^+$T cells presence, we assume that the suppression from the tumor is later than the CD8$^+$T cells proliferation.[4] Correspondingly, we set a larger value for $u$ than for $k_1$ as $1e^5$.

Assumption 3: The maximum cytotoxic drugs' effect on upregulating checkpoint presentation, $p$, is set to be 0.2.

Assumption 4: In the literature, the decay rate of cytotoxic drug ($\gamma_M$) is assumed to be 0.9. So, we took the decay rate $\gamma_I$ as 0.9 , too.[5]

Assumption 5: In the literature, chemotherapy's killing effects on tumor cells and CD8$^+$T cells were assumed to be 0.9 and 0.6. In this paper, we adjust them to be 0.6 and 0.6.[5]

**Extended Table 1.** Parameters in TIME dynamical evolutionary model

| Parameter | Description | Estimated value |
|---|---|---|
| $T_{max}$ | carry capacity of tumor cells | $2.6e^{8}$ |
| $\alpha_{sr}, \alpha_{rs}$ | competition coefficients between sensitive and resistant tumor cells | 0.7,0.9 |
| $m_L$ | CD8+T cells death rate | 0.05 |
| $T_{max}$ | carry capacity of CD8$^+$T cells | $1e^{7}$ |
| $d$ | killing power of CD8$^+$T cells | 2.34 |
| $s$ | Michaelis constant of tumor lysed by CD8$^+$T cells | $8.39e^{-2}$ |
| $u$ | Michaelis constant of tumor's effects on accumulating regulatory T cells | $1e^{5}$ |
| $k_1, k_2, k_3$ | Michaelis constant of antigens simulating CD8$^+$T cells' proliferation | $1e^{4}$ |
| $p$ | maximum cytotoxic drugs' effect on upregulating checkpoints presentation | 0.2 |
| $\gamma_I$ | decay rate of immune checkpoint inhibitors | 0.9 |
| $\gamma_M$ | decay rate of cytotoxic drug | 0.9 |
| $K_{Ts}$ | cytotoxic drug's killing effects on sensitive tumor cells | 0.6 |
| $K_L$ | cytotoxic drug's killing effects on CD8$^+$T cells | 0.6 |

#### 1.3. Data fitting

##### 1.3.1. Biomarkers preprocessing

PD-1-associated gene is PDCD1. Because the distribution of PDCD1 expression ($\rho$) is long tail, we use log normalization. The fraction of CD8$^+$T cells suppressed by immune checkpoints is assumed to be $\mu_0 = \frac{\log(\rho+1)}{\log(\max(\rho)+1)}$.

Antigen presentation molecules associated genes are MHC-I and MHC-II associated HLA genes. After applying ssGSEA, we can get the ssGSEA score. We normalize this score by Min-Max normalization.

The ratio of regulatory T cells to CD8+T cells can be derived directly from the CIBERSORT analysis result.

The distribution of CD8+T cell signatures is close to a normal distribution. We want to convert the CD8$^+$T cell signatures to the CD8$^+$T cell infiltration ($\eta_{LT}$). Because in our model, when the ratio of CD8$^+$T cells to tumor cells $\eta_{LT}$ equals to 0.03, the CD8$^+$T cells killing tumor term, $d\left(\frac{L(1-\mu)}{T}\right)^l / \left(s + \frac{L(1-\mu)}{T}\right)^l$ equals 0.025, which is more significant than non-metastatic melanoma intrinsic growth rate, contrary to the fact that the tumor is successfully immune escape. Consequently, we assume that maximum $\eta_{LT}$ equals 0.03 before treatment interaction. Then, the CD8$^+$T cells signature distribution can be converted into corresponding value $\eta_{LT}$, ranging from 0 to 0.03.

#### 1.3.2. Estimation of parameters without treatment

As we mentioned in the main manuscript, the unknown parameters without treatment that we want to estimate are regulatory effects from tumor cells and regulatory T cells on CD8$^+$T cells $(q, h)$ and association index of MHC-I and MHC-II expression on antigen presence on tumor surface $(\omega)$. Simultaneously, we want to estimate the latent variables $z_i$, patient-specific antigens process ability ($r$). We adopt the idea of an estimation maximization algorithm to design our parameters estimation algorithm. The EM algorithm can be viewed as two alternating maximization steps. That is, at the E step and M steps, latent variables and unknown parameters are calculated to maximize the given criteria. Our algorithm is a similar iterative algorithm.

Step 1: Initialize the value of latent variables, patient-specific antigens process ability ($r$), as 0.1.
Step 2: Set the initial cells population of $T_s,\ T_r,\ L$ to be $1e^4, 4e^1, 1e^2$, randomly assign tumor intrinsic growth rate $a_s$ and $a_r$, and input the observable patient-specific variables $(x_i)$: the expression of MHC-I and MHC-II molecules (measured by the ssGSEA score $\phi$, a normalized summation of both molecules) and the ratio of regulatory T cells to CD8$^+$T cells $(\eta_{RL})$. Search for the best estimate of unknown parameters to minimize the mean absolute error between the predicted ratio of CD8+T cell population to tumor cell population and the estimated ratio by CD8$^+$T cell signatures expression.
Step 3: Compute the better estimate of patient-specific antigen process ability (r) to minimize the mean absolute error defined in step 2.
Step 4: Iterate steps 2 and 3 until convergence.

The search range of $q, h$ and $\omega$ are (0.01,0.02,0.03), (0.01,0.02,0.03), and (2,4,6,8), respectively. The best estimate pair of $q, h$ and $\omega$ are 0.03, 0.01, 6. The estimated repressive effects directly from tumors is larger than those from regulatory T cells. The final mean absolute error of the simulated ratio of CD8+T cells to tumor cells and the actual ratio is 0.003. The relative error is about 0.25.

#### 1.3.3. Estimation of parameters associated with ICI treatment

The unknown parameter related to ICI treatment is the rate of antigen presence by CD8$^+$T cells induced immunogenetic tumor death $(j_L)$. We estimate this parameter in the following step:

Step 1: Initialize cells population of $T_s,\ T_r,\ L$ to be $1e^4, 4e^1, 1e^2$, randomly assign tumor intrinsic growth rate $a_s$ and $a_r$ and set the value of $q, h$ and $\omega$ to be 0.03, 0.01 and 6. Then, estimate the specific antigens process ability ($r$) for patients in the Liu dataset to minimize the mean absolute error between the predicted ratio of CD8+T cell population to tumor cell population and the estimated ratio by CD8$^+$T cell signatures expression.
Step 2: Simulate the ICI treatment by our ODE model and find the best estimate of $j_L$

to predict patients' response results.

The estimated value of $j_L$ is 0.11.

**Extended Table 2.** Estimated key parameters

| Parameter | Description | Estimated value | Source |
|---|---|---|---|
| $q$ | Tumor cells suppressive effects on $CD8^+T$ cells | 0.03 | TCGA SKCM |
| $h$ | Regulatory T cells suppressive effects on $CD8^+T$ cells | 0.01 | TCGA SKCM |
| $\omega$ | Parameter of ssGSEA score of MHC-I and MHC-II on $j_c$ | 6 | TCGA SKCM |
| $j_L$ | Antigen presence rate by $CD8^+T$ cells induced immunogenetic tumor death | 0.11 | Liu |

The chemotherapy treatment efficacy is affected by tumors' heterogeneous chemotherapy resistance. This information has not been recorded in the TCGA SKCM dataset, so it is a pity that we cannot derive parameters related to chemotherapy from the dataset. We assume the value of antigen presence by chemotherapy-induced immunogenetic tumor death $(j_M)$ is 30 percent of $CD8^+T$ cells induced $(j_L)$, 0.033. In our model, if the value of $j_M$ is too high, chemotherapy can induce enough $CD8^+T$ cell proliferation, substituting the usage of ICI, which is contrary to reality.

**Supplementary reward shaping for ICC decision**

**2.1 ICI cost ($\lambda_1$)**

ICI can help CD8+T cells eliminate the tumor, the terminal reward for tumor elimination (equals 1). So, we want to set the ICI cost smaller than 1 to lead the agent using ICI to eliminate tumors. Because the maximum cycles for ICI usage are 10, we set the ICI cost $\lambda_1$ every cycle as 0.04, making the maximum total ICI cost equal to 0.4, smaller than 1.

**2.2 Early tumor elimination reward**

$\lambda_2(4 - T)$ is an additional term rewarding the early tumor elimination before the fourth cycle. We set $\lambda_2$ equal to 0.1.

**Supplementary classification of extremely cold, cold, and hot tumors**

**3.1 Classification rules**

We classified the TIME of patients into three categories based on their ratio of $CD8^+T$ cell population to tumor cell population ($\eta_{LT}$ ) and $CD8^+T$ cell growth rate so that their DRL-derived schedules and trajectories can be explicitly clustered (Figure 4b). We denote tumors whose CD8+T cell population grows quite fast and can be effectively stimulated by low dosages of chemotherapy as "hot tumors". We summarize that

their $CD8^+T$ cell net growth rate $rj_C - \ (m_L + \eta_{RL}h + q)$ is larger than 1.2 times tumor growth rate. We denote tumors whose CD8+T cell infiltration is extremely low and unlikely to be turned hot as “extremely cold tumors”. We summarize that their ratio of $CD8^{+}$T cell population to tumor cell population $\eta_{LT}$ is lower than 0.006. The remaining tumors between these two criteria are denoted as “cold tumors”.

### REFERENCES


1. Zhang J, Cunningham JJ, Brown JS, Gatenby RA. Integrating evolutionary dynamics into treatment of metastatic castrate-resistant prostate cancer. Nature Communications. 2017;8(1):1816.
2. Zhang J, Cunningham JJ, Brown JS, Gatenby RA. Integrating evolutionary dynamics into treatment of metastatic castrate-resistant prostate cancer. Nature Communications. 2017;8(1):1816.
3. Ribeiro RM, Mohri H, Ho DD, Perelson AS. In vivo dynamics of T cell activation, proliferation, and death in HIV-1 infection: why are CD4+ but not CD8+ T cells depleted? Proceedings of the National Academy of Sciences. 2002;99(24):15572–7.
4. Liu YT, Sun ZJ. Turning cold tumors into hot tumors by improving T-cell infiltration. Theranostics. 2021;11(11):5365.
5. de Pillis LG, Gu W, Radunskaya AE. Mixed immunotherapy and chemotherapy of tumors: modeling, applications and biological interpretations. Journal of Theoretical Biology. 2006;238(4):841–62.
